# Surface changes of clinical target volume and its sublocations in nasopharyngeal cancer with image-guided radiation therapy

**DOI:** 10.1101/2022.05.15.22275068

**Authors:** Wenyong Tan, Ming Yang, Desheng Hu

## Abstract

**PURPOSE:** We aimed to quantify the sublocal geometric uncertainties of the neck prophylactic clinical target volume (CTV_prophy_) during image-guided radiotherapy for nasopharyngeal cancer (NPC).

**MATERIALS AND METHODS:** Twenty patients with locally advanced NPC underwent one planning computed tomography (CT_plan_) followed by six weekly CT (CT_repeat_) scans during chemotherapy and intensity-modulated radiation therapy. The sternocleidomastoid muscle (SCM) and its anterior, middle, and posterior parts, as well as the body contours at the 1^st^ (C1) and 2^nd^ (C2) cervical vertebrae, hyoid bone (HB), and cricoid cartilage (CC) in transverse CT sections, were manually delineated in the CT_plan_ and each CT_repeat_. The residual error and 2D or 3D vector displacements of each sublocation were calculated, and the planning target volume (PTV) margins were estimated using the PTV margin formula.

**RESULTS:** The left- and right-sided SCM volume decreased by 3.7 ± 9.6% (1.9–5.4%) and 5.1 ± 6.7% (3.9–6.3%), respectively, and the center of mass shifted medially 0.8–0.9 mm. An anisotropic PTV margin of 2–4 and 1–5 mm was needed in the left-right and anterior-posterior directions, respectively. The geometric changes in the upper neck at the C1 and C2 sections were smaller than those in the middle-lower neck at the HB and CC levels. At the same sublocation, the margin needed in the anterior-middle part was smaller than that needed in the posterior part of the neck. The rigid imaging registration-induced anatomical errors in the upper neck were < 1.9%, and those in the middle and lower neck level were 0.6–3.8%.

**CONCLUSIONS:** The surface geometrical changes of the neck prophylactic CTV in the sublocations are substantial and an anisotropic PTV margin of 1–5 mm is needed in the context of image-guided radiotherapy for NPC.

## INTRODUCTION

Intensity-modulated radiation therapy (IMRT) for head and neck cancer (HNC) has been shown to improve local control and survival outcomes, as well as decrease toxicities[1-3]. To achieve desired treatment outcomes from IMRT, accurate definitions of the target volume and organs at risk (OARs) are vital. Initial delineations from planning computed tomography (CT) have been used in IMRT planning and execution [4]. Additionally, the target volumes [5-10], OARs [11, 12], and sublocations [9, 13] displayed considerable changes in tumor response to radiation treatment and weight loss. Adaptive IMRT, including re-delineation [14], is a feasible solution for sparing the OAR without compromising the radiation dose to the target volume [15]. Target volume delineation in planning CT and adaptive target volume delineation has been based on various guidelines [16-18], in which the definitions of neck lymph node levels and related clinical target volumes (CTVs) were mainly based on the anatomical boundaries in transverse CT sections such as the sternocleidomastoid muscle (SCM), 1^st^ (C1) and 2^nd^ (C2) cervical vertebra, hyoid bone (HB), and cricoid cartilage (CC). However, these anatomical structures, especially the SCM, result in considerable geometrical changes during IMRT which may differ among each neck sublocation. In this study, we assumed that the geometrical changes of the anatomical structures in the planning CT were identical to the surface of the neck prophylactic CTV (CTV_prophy_). The aims of this study were to quantify the geometrical changes of the selected anatomical structures, including the sublocations of the SCM during chemoradiotherapy for nasopharyngeal cancer (NPC), and to estimate the planning target volume (PTV) margin at various CTV_prophy_ sublocations. These findings will be valuable for improving adaptive radiation treatment for NPC.

## MATERIAL AND METHODS

### Image data and registration

Twenty patients with locally advanced NPC (stage III–IVb based on the American Joint Cancer Committee criteria from 2011) who received IMRT and chemotherapy were recruited. This study was approved by the ethics committee of Hubei Cancer Hospital, Wuhan, China [7]. Thirteen patients received concurrent chemotherapy with platin and seven underwent sequential therapy. Each patient underwent one planning CT scan (CT_plan_) with intravenous contrast and six weekly repeat CT scans (CT_repeat_) without contrast [7, 10]. The patients wore a five-pointed thermoplastic mask covering the head, neck, and shoulders during the repeat and planning CT scans. The CT_repeat_ images were acquired at every five fractions and exported as DICOM digital data. They were then imported into the research software developed by Department of Radiation Oncology, The Netherlands Cancer Institute, Amsterdam, The Netherlands [7, 19]. Each CT_repeat_ was rigidly registered with the respective CT_plan_ using bone matching, and the matching areas were large enough to cover the entire potential PTV [7]. The aim of this registration was to make all images of one patient share the same three-dimensional coordinates to calculate the geometrical changes, which was performed according to our standard operating procedure of image-guided IMRT. None of the IMRT procedures were modified with the aim of documenting the geometrical variations of the sublocations during the course of IMRT.

### Definitions of anatomical reference

In IMRT for HNC, several neck nodal levels of the CTV_prophy_ and most of the anatomical boundaries are based on sublocations such as the SCM, C1, C2, HB, and CC, as they can be unambiguously defined on transverse CT sections to minimize inter-observer uncertainties [16-18]. For example, the deep surface of the SCM makes up the lateral boundaries of level II and III as well as the anterior surface of level IVa and IVb. Meanwhile, the anterior edge of the SCM makes up the anterior boundaries of level III and IVa as well as the posterior boundary of level VIII and the lateral boundary of level VIa. The posterior edge of the SCM makes up the posterior boundaries of level II, III, and IVa; and the anterior edge of level V and Xb; as well as the posterior edge of level Xa. Additionally, the caudal edge of the lateral process of C1 comprises the cranial edge of level II, and the caudal edge of the HB makes up the caudal edge of levels Ib and II and the cranial edge of level III. Finally, the CC makes up the caudal edge of level II and the cranial edge of level IVa [16].

Anatomically, the SCM, as the largest and most superficial paired cervical muscle, originates at the manubrium of the sternum and clavicle, and extends posterior-cranially into the whole neck region with an insertion at the mastoid process of the temporal bone of the skull [20]. This muscle is 14–18 cm long and 3–5 cm wide in the middle [20]. The neck provides mobility and stability to the head [20], which suggests that considerable geometric variability in the neck sublocations may be present in head and neck IMRT. Based on the cervical vertebrae, the neck can be anatomically divided into the upper part, which includes all anatomical structures at the C1 and C2 sections, and the middle and lower parts including those from C3–C7 [20].

To quantify the geometric variability of the neck sublocations, we manually contoured the SCM in all CT images and arbitrarily selected the caudal edge of the lateral process of C1 (C1S), the cranial (C2S) and caudal (C2I) edges of C2, and the caudal edge of the HB and CC on various axial CT sections (Fig 1A). For each of the five axial CT sections, we manually contoured the anterior, middle, and posterior points at the medial edge of each SCM. Each sublocation was contoured as the region of interest (ROI) in the shape of a 3 mm × 3 mm square (Fig 1B). To minimize the uncertainty in the definition of the ROI, the center of the ROI exactly co-located the most anterior-posterior edge, as well as the middle point between the anterior and posterior located at the medial edge of each SCM (Fig 1B). Because all ROIs were defined by the anatomical boundaries of various neck nodal levels [16], we could safely assume that the geometrical variations of these ROIs accurately represented the sublocation changes of the CTVprophy surface during the course of radiation treatment.

**Figure 1.**
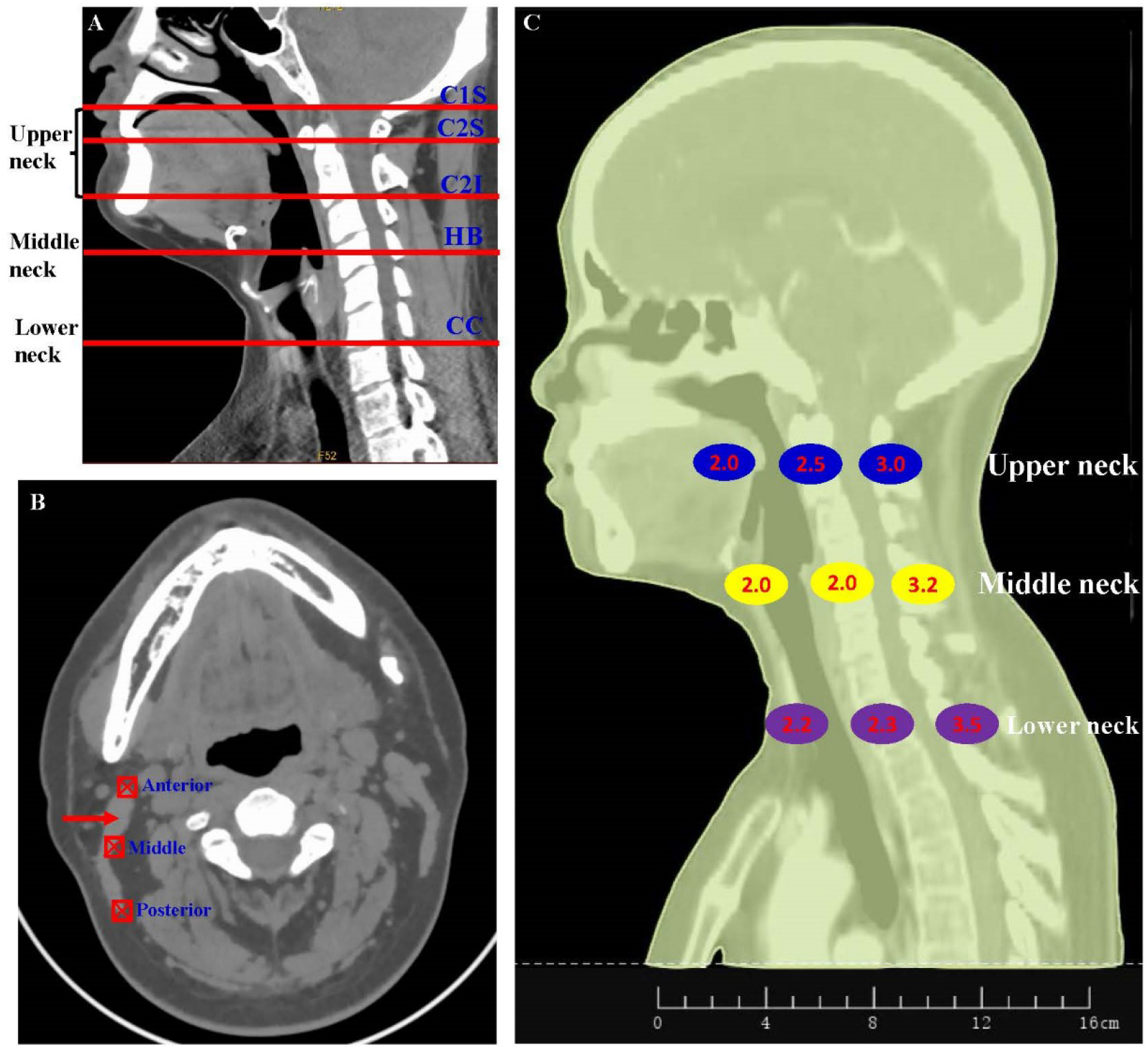
The sublcocations at the upper, middle, and lower neck located at the caudal edge of the lateral process of C1 (C1S), C2 (C2I), the hyoid bone (HB), and the cricoid cartilage (CC), as well as the cranial edge of C2 (C2S). C1S, C2S, and C2I represent the upper neck, while HB and CC represent the middle and lower neck respectively (A). The anterior, middle, and posterior edge of the sternocleidomastoid muscle (SCM) were at the lateral edge of the prophylactic CTV and represent the CTV surface. The centers of the red squares are collocated at the edge of the SCM, and the red arrow indicates the SCM (B). The geometric changes were quantitively estimated in various neck sublocations (C). SCM_L and SCM_R were the left- and right-sided SCM respectively. The lines show the means, and the error bars show the 95% confidence intervals.

Between the CT_repeat_ and CT_plan_, the upper neck could accurately match due to the substantial rigid bones in the head; however, the middle and lower parts of the neck could not accurately match due to significant deformations in the neck, which could induce registration errors (error_reg_). To quantify the sublocation error_reg_, we manually delineated the body contour on the C1, C2, HB, and CC sections of the axial CT slice in the registered (Contour__reg_) and anatomically real imaging sections (Contour__real_). The error_reg_ was calculated as 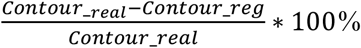. We also manually delineated the external contours of the whole head and neck superior to the cranial edge of the sternal manubrium and those at C1S, C2S, C2I, HB, and CC in the axial CT section. The whole and sublocal external contours could be automatically calculated as the volume and area, respectively. The sublocal geometric changes and error_reg_ were qualified according to previous studies [7, 9]. To minimize inter-observer variability, all ROI definitions were performed by a single radiation oncologist (W.T.) devoted to HNC.

### Calculation of geometrical variation and estimation of PTV margin

For each ROI, the volume, center-of-mass (COM) position, and area of the external contour in various sublocations were automatically calculated [7, 19]. For the global SCM, the position displacement in the left-right (LR), anterior-posterior (AP), and cranial-caudal (CC) directions could then be calculated manually. For the ROIs of the sublocations of the SCM, the positional displacement could only be calculated in the LR and AP directions, as CC displacement could not be easily quantified due to the use of only one axial CT slice. For each direction of the ROIs, six displacements were used to analyze the group mean (M) and systemic (∑) and random (σ) error [4, 7] as well as the 3D or 2D vector displacement [7]. Using the volume and area parameters in the CT_plan_ as the reference, the volume loss of the SCM and area changes of the external contour in C1S, C2S, C2I, HB, and CC were calculated and used to present the global and sublocal variations. The van Herk margin formula (2.5∑ + 0.7σ) [4, 21] was used to calculate the PTV margin in each direction of all CTV_prophy_ sublocations.

### Statistical analyses

An independent t test was used to compare the SCM volume difference between the left and right side, as well as the contour_reg_ and contour_real_. The SCM volume among the different weeks was analyzed using one-way analysis of variance. The correlation between patient weight loss and SCM volume was estimated using univariate linear regression analysis. All tests were two-tailed and a 5% significance level was used to establish statistical significance. Statistical Package for the Social Sciences (SPSS version 24.0; IBM Corporation, Armonk, NY, USA) and Microsoft Office Excel (Microsoft Office 2016; Microsoft Corporation, Redmond, WA, USA) were used for all statistical analyses.

## RESULTS

### Global changes of SCM

The volume of the SCMs in both sides showed a linearly decreasing trend throughout the treatment course (Fig. 2A). The average volume was reduced by 3.7±9.6% (95% CI: 1.9–5.4%) and 5.1±6.7% (3.9–6.3%) in the left- and right-sided SCM, respectively, and there was no statistical difference (*P* = 0.195). Moreover, no significant difference was noted among the various weeks (*P* > 0.05). For both sides of the SCM, patient weight loss was significantly related to SCM volume reduction (Fig. 2B and 2C).

**Figure 2.**
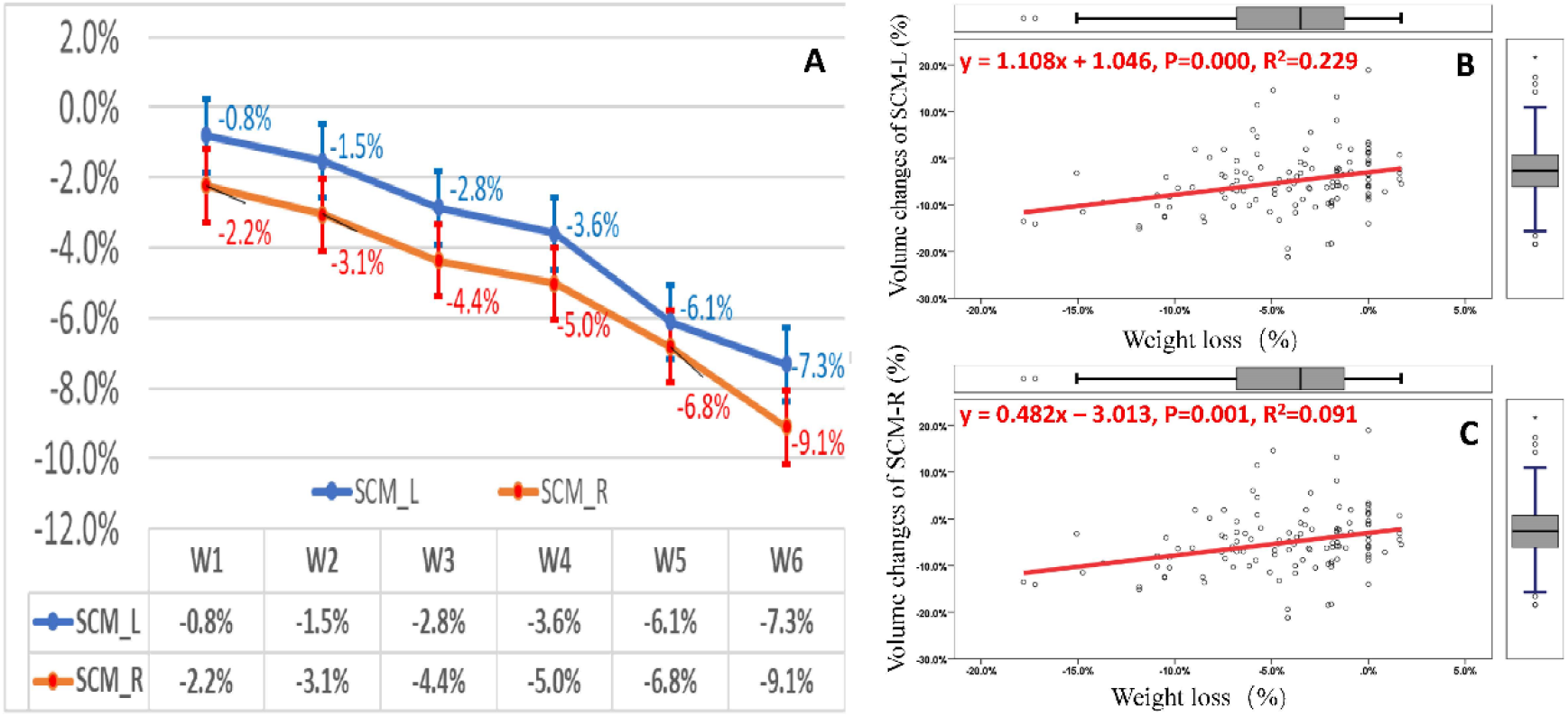
Both sides of the SCM demonstrated decreasing volume with time (A). The volume loss was significantly related to weight loss in NPC patients (B, C). The lines show the means, and the error bars show the 95% confidence intervals.

The global position displacements of both sides of the SCMs are listed in Table 1. Medial shifts of 0.8–0.9 mm and caudal shifts of 0.5 mm were observed. The systemic error in all three directions was < 0.6 mm and the random error was >1.0 mm. The 2D vector displacements of both sides of the SCMs were 3.0–3.5 mm and the PTV margins needed were of 1.7–2.3 mm in the LR and CC directions and ≤ 1 mm in the AP direction as shown in Table 1.

**Table 1.** Global position variations of SCMs and the estimated PTV margin (mm)

|  | Left |  |  |  | Right |  |  |  |
| --- | --- | --- | --- | --- | --- | --- | --- | --- |
| Direction | M | $\Sigma$ | $\sigma$ | Margin* | M | $\Sigma$ | $\sigma$ | Margin |
| Left-right | -0.9<sup>#</sup> | 0.4 | 1.1 | 1.8 | 0.8 | 0.4 | 1.1 | 1.8 |
| Cranial-caudal | -0.5<sup>#</sup> | 0.6 | 1.1 | 2.3 | -0.5<sup>#</sup> | 0.4 | 1.0 | 1.7 |
| Anterior-posterior | 0.01 | 0.03 | 1.0 | 0.8 | 0.01 | 0.06 | 1.2 | 1.0 |
| 3D vector displacement<sup>§</sup> | $3.5 \pm 1.6$ (1.1, 6.1) | | | | $3.0 \pm 1.8$ (0.6, 8.5) | | | |
| Volume reduction<sup>§</sup> | $3.7\% \pm 9.6\%$ (1.9, 5.4%) | | | | $5.1\% \pm 6.7\%$ (3.9, 6.3%) | | | |

Abbreviations: M, average displacement; $\Sigma$ , systemic error; $\sigma$ , random error; SCM, sternocleidomastoid muscle; PTV, planning target volume
\*The margin was estimated using van Herk's formula: $2.5\Sigma+0.7\sigma$ .
#Negative numbers represent movement to the right, posterior, or caudal direction.
§Data are shown as the mean ± standard deviation, and the 95% confidence intervals are in brackets.

### Positional shifts in SCM sublocations

The position displacements in the axial C1 and C2 sections were replaced with the upper neck, and the HB and CC sections were replaced with the middle and lower neck, respectively. In the LR direction, all SCM sublocations moved medially, with a group mean of 0.4–1.7 mm. For the upper neck, the anterior and middle SCMs shifted medially by 0.4–1.7 mm and those in the posterior part of the SCM shifted 0.4–1.0 mm as shown in Table e1. For the middle and lower neck, the displacements in the anterior and middle part were 0.1–0.8 mm and those in the posterior part were 0.2–1.0 mm. The systemic errors were < 1.0 mm in all directions, and the random errors were ≥ 1.0 mm (Table 2). The PTV margins in the LR direction were 1.6–2.6 mm (Table 2) and 2.3–3.6 mm in the anterior-middle and posterior part of the SCM, respectively (Table 3).

**Table 2.** Position shifts and recommended margins in the left-right direction at the various axial sites of SCM(mm)

| Sublocations | Left-sided SCM |  |  |  | Right-sided SCM |  |  |  |
| --- | --- | --- | --- | --- | --- | --- | --- | --- |
| | M | $\Sigma$ | $\sigma$ | Margin* | M | $\Sigma$ | $\sigma$ | Margin |
| Cranial edge of C2 |  |  |  |  |  |  |  |  |
| Anterior<sup>#</sup> | -0.7<sup>§</sup> | 0.4 | 0.8 | 1.6 | 0.4 | 0.5 | 1.0 | 2.0 |
| Middle<sup>#</sup> | -0.8 | 0.6 | 1.3 | 2.4 | 1.0 | 0.5 | 1.3 | 2.2 |
| Posterior<sup>#</sup> | -0.4 | 0.8 | 1.7 | 3.2 | 0.5 | 0.9 | 1.9 | 3.6 |
| Caudal edge of C2 |  |  |  |  |  |  |  |  |
| Anterior | -1.7 | 0.6 | 1.1 | 2.3 | 1.2 | 0.6 | 1.0 | 2.2 |
| Middle | -1.5 | 0.6 | 1.6 | 2.6 | 1.4 | 0.6 | 1.5 | 2.6 |
| Posterior | -0.7 | 0.6 | 1.9 | 2.8 | 1.0 | 0.7 | 1.9 | 3.1 |
| Caudal edge of hypoid bone |  |  |  |  |  |  |  |  |
| Anterior | -0.6 | 0.3 | 0.8 | 1.3 | 0.4 | 0.4 | 0.9 | 1.6 |
| Middle | -0.4 | 0.4 | 1.1 | 1.8 | 0.8 | 0.4 | 0.9 | 1.6 |
| Posterior | -0.2 | 0.5 | 1.5 | 2.3 | 1.0 | 0.5 | 1.6 | 2.4 |
| Cranial edge of cricoid cartilage |  |  |  |  |  |  |  |  |
| Anterior | -0.5 | 0.6 | 1.0 | 2.2 | 0.3 | 0.5 | 1.4 | 2.2 |
| Middle | -0.1 | 0.4 | 1.1 | 1.8 | 0.3 | 0.5 | 1.1 | 2.0 |
| Posterior | -0.4 | 0.8 | 1.7 | 3.2 | 0.7 | 0.7 | 1.7 | 2.9 |

Abbreviations: SCM, sternoclavicular muscle; M, group mean; $\Sigma$ , systemic error; $\sigma$ ,
random error; C2, 2nd cervical vertebrae
\*The margin was estimated using van Herk's formula: $2.5\Sigma+0.7\sigma$ .

**Table 3.** Position shifts and recommended margins in the anterior-posterior direction at the various axial sites of SCM (mm)

| Sublocations | Left-sided SCM |  |  |  | Right-sided SCM |  |  |  |
| --- | --- | --- | --- | --- | --- | --- | --- | --- |
| | M | $\Sigma$ | $\sigma$ | Margin* | M | $\Sigma$ | $\sigma$ | Margin |
| Cranial edge of C2 |  |  |  |  |  |  |  |  |
| Anterior<sup>#</sup> | 0.2 | 0.3 | 0.5 | 1.1 | 0.1 | 0.4 | 0.6 | 1.4 |
| Middle<sup>#</sup> | 0.1 | 0.5 | 0.6 | 1.7 | 0.4 | 0.2 | 0.4 | 0.8 |
| Posterior<sup>#</sup> | 0.2 | 0.5 | 0.9 | 1.9 | 0.1 | 0.4 | 0.9 | 1.6 |
| Caudal edge of C2 |  |  |  |  |  |  |  |  |
| Anterior | 0.1 | 0.4 | 0.7 | 1.5 | -0.1<sup>§</sup> | 0.3 | 0.7 | 1.2 |
| Middle | 0.5 | 0.3 | 0.5 | 1.1 | 0.4 | 0.7 | 0.9 | 2.4 |
| Posterior | 0.8 | 0.3 | 0.7 | 1.2 | 0.4 | 0.4 | 0.9 | 1.6 |
| Caudal edge of hyoid bone |  |  |  |  |  |  |  |  |
| Anterior | -0.2 | 0.2 | 0.5 | 0.9 | -0.4 | 0.5 | 0.9 | 1.9 |
| Middle | -0.2 | 0.3 | 0.6 | 1.2 | -0.1 | 0.4 | 0.8 | 1.6 |
| Posterior | 0.9 | 0.4 | 1.0 | 1.7 | 0.6 | 0.7 | 1.0 | 2.5 |
| Cranial edge of cricoid cartilage |  |  |  |  |  |  |  |  |
| Anterior | -0.3 | 0.5 | 0.9 | 1.9 | -0.1 | 0.8 | 1.3 | 2.9 |
| Middle | -0.7 | 0.5 | 1.0 | 2.0 | -0.2 | 0.8 | 1.3 | 2.9 |
| Posterior | -0.5 | 1.2 | 2.0 | 4.4 | -0.3 | 1.0 | 1.8 | 3.8 |

Abbreviations: SCM, sternocleidomastoid muscle; M, group mean; $\Sigma$ , systemic error; $\sigma$ , random error; C2, 2nd cervical vertebrae
\*The margin was estimated using van Herk's formula: $2.5\Sigma+0.7\sigma$ .
#The anterior, middle, and posterior site of SCM
§Negative numbers represent movement to the right, posterior, or caudal direction.

In the AP direction, the position in the upper neck shifted anteriorly 0.1–0.5 mm in the anterior-middle part of both SCMs and posteriorly 0.1–0.7 mm in the middle and lower neck. The posterior part of the SCMs shifted anteriorly 0.1–0.9 mm in the upper and middle neck and posteriorly 0.3–0.5 mm. The systemic and random errors in all directions were <1.0 mm in the upper and middle neck. However, in the lower neck, the systemic errors were 0.5–1.2 mm and the random errors were 0.9–2.0 mm (Table 3). In the AP direction, the PTVs were 0.8–2.5 mm and 1.9–4.4 mm in the upper-middle and lower neck, respectively (Table 3).

The 2D vector displacements at various sublocations of the SCMs were heterogeneous at 1.6– 2.8, 2.2–3.1, and 2.9–3.7 mm in the upper, middle, and lower neck, respectively. Finally, the 2D displacements were 1.6–3.7 mm and 1.9–3.2 mm in the left-and right-side, respectively, as shown in Table 4.

**Table 4.**
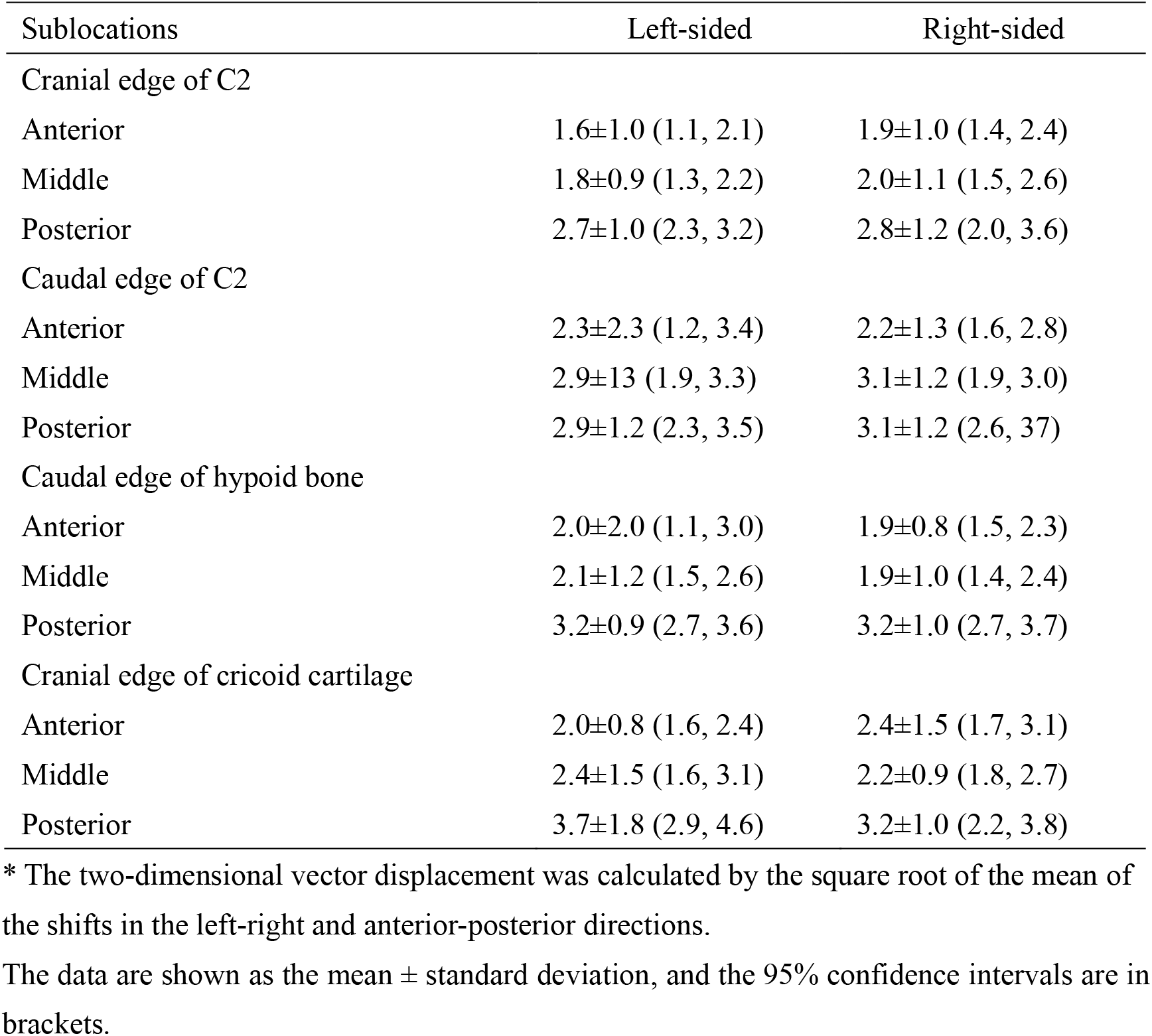
Two-dimensional vector displacements^*^ of the internal surface at various axial sites of SCM (mm)

### Registration induced errors in sublocations

In the C1, C2, HB, and CC axial sections, the areas of the local external contour showed decreasing time trends throughout the treatment course (Fig. 3A and 3B). For the upper neck, the mean area reduction of the external contour was 3.4–4.5% (Fig. 3A), and that of the middle and lower neck was 4.8–7.8% (Fig. 3B). As the error_reg_, nearly no difference between the contour_reg_ and contour_real_ was observed. For the entire external contour, the contour_reg_ and contour_real_ decreased by 6.1 ± 1.5% (0.5–12.1%) and 5.9 ± 6.3% (0.6–16.3%), respectively (Fig. 3C). The error_reg_ of the entire external contour was 0.2 ± 6.3% (−10.0–12.0%), as shown in Fig. 3D.

**Figure 3.**
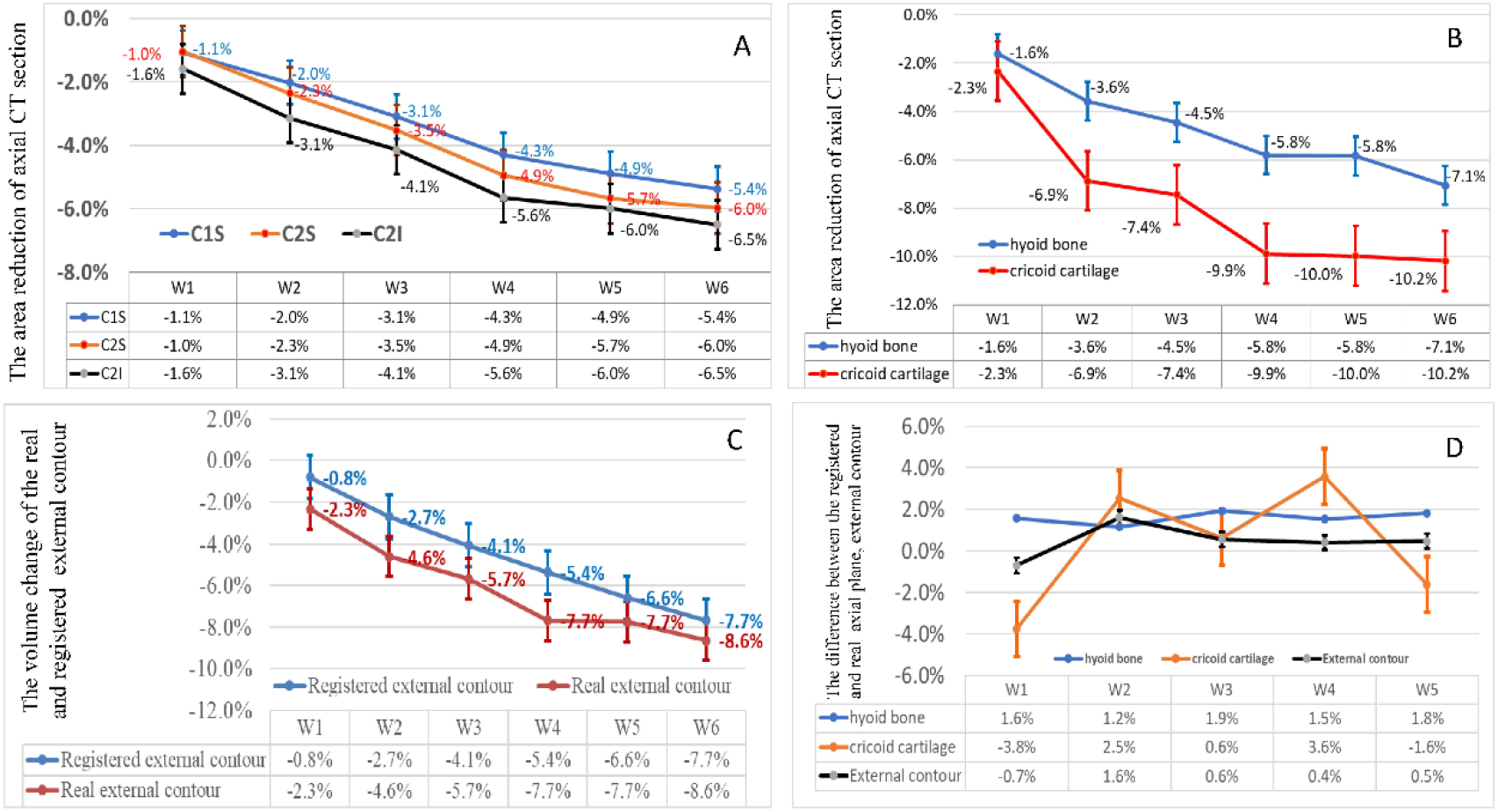
The transverse external contour of various sublocations in the upper (A), middle and lower (B) neck, as well as the registered and real external contour (C) decreased as radiation fractions elapsed. And the registration-induced imaging errors in the middle and lower neck as well as the neck body contour were demonstrated at different treatment time (D). The lines show the means, and the error bars show the 95% confidence intervals.

For the middle and lower neck, the error_reg_ as sublocal uncertainty was 1.7 ± 8.4% (−7.5–26.3%) and -0.4 ± 19.8%(−33.0–31.3%), respectively (Fig. 3D). Thus, the lower neck in the CC section showed much larger local geometric variation. All the area reductions in the contour_reg_ correlated significantly with weight loss, while those in the contour_real_ did not.

## DISCUSSION

For HNC, the primary tumor and positive lymph nodes in the neck usually demonstrate substantial shrinkage, and their surrounding healthy organs may undergo considerable geometric changes over the course of radiation treatment [5-7]. This study, provided more nuanced understanding of the geometrical variation of neck sublocations and estimated their margins to establish the planning target volume. Generally, geometrical changes are larger in the lower and posterior parts of the neck than in the upper and anterior parts. As the anatomical structure used to define the prophylactic CTV at the neck nodal levels, the SCM shifted medially <1 mm. In most neck sublocations, an anisotropic margin of 1–5 mm is needed to establish the PTV from the elective CTV in most directions; however, in the posterior part of the lower neck, this number is larger.

### Geometrical changes in head and neck cancer

Image-guided IMRT with steep dose gradients enables accurate radiation delivery to tumors [22]. However, during IMRT, geometrical changes in the tumors and/or OARs will most likely result in uncertainties in the dose distribution. Previous studies have shown that at the end of IMRT, the mean gross tumor volume (GTV) decreased by 55.3–70.0% and the CTV by 52.0– 71.1% [5, 7]. Additionally, the volume loss rates per treatment day were found to be 1.8–3.9% for the GTV and 0.4–2.3% for the CTV [5-7]. Similarly, the center of mass of the GTV and CTV position shifts were 0.5–1.3 mm [6, 7] and 1.5–3.1 mm [6], respectively. Furthermore, the prophylactic CTV, which usually included several neck nodal levels, also demonstrated considerable volume and position changes. Specifically, the volume loss was 0.3–0.5%, and the COM displacement was 0.5–1.8 mm per treatment day [6, 7]. These studies confirmed that both the volume and position of the target volumes change substantially. In this study, we showed that the volume of the SCM decreased by 4–5% on average and shifted medially by nearly 1 mm.

Volume loss and position changes of the target volume induce uncertainty throughout the course of IMRT. In the previous study, the Dice similarity index of all target volumes between the planning and repeat CTs for NPC was <0.7 after the third week, suggesting that the volume coverage after three weeks was inadequate. Thus, adaptive replanning is a possible solution for minimizing the uncertainties from geometrical changes.

### Margin from CTV to PTV

Usually, to establish the PTV, a safety margin is expanded from the CTV to ensure that an adequate radiation dose is administered. However, setup and organ position uncertainties may emerge during the treatment process. For HNC, the CTV-PTV margin is usually 3–5 mm [8, 23, 24] and the head motion within the mask is estimated to be approximately 1–3 mm [25]. Additionally, image-guided technology may reduce geometrical uncertainties; requiring a smaller margin [26], maybe as low as <5 mm, to reduce irradiation to extra tissue [27]. The combination of adaptive replanning and margin reduction might spare the OARs, particularly the parotid glands [28]. Generally, conventional isotropic margins may be suboptimal. Therefore, with advanced imaging guidance, the PTV may be anisotropic to reduce normal tissue exposure without compromising target coverage [29]. In this study, we used repeat CT as offline imaging guidance to mimic daily IMRT delivery. We suggest an anisotropic margin of 1–5 mm in various neck sublocations.

The neck is a complex site for the definition of prophylactic CTV and requires geometric accuracy. With the rigid registration of bony matches, sublocation setup errors may exceed the residual global patient setup errors and, as a result, the overall setup accuracy may overestimate the radiation treatment precision [30]. The overall displacement was 0.1–0.5 mm and the systemic error in the three directions were 1.1–1.2 mm. The largest systemic errors were found for the mandible, larynx, occiput bone, and the caudal edge of C7 at 2.2–2.4 mm [30]. A PTV margin of 5 mm was adequate to cover the global patient setup uncertainty, however, it might not be sufficient for some sublocations [30]. The tumor shape variability in oropharyngeal cancer depends on the tumor sublocation and volume. The cranial and caudal borders in the posterior pharyngeal wall are at high risk of insufficient coverage during IMRT [31]. In this study, the geometrical uncertainties of several edges of prophylactic nodal levels also depended on their sublocations. Furthermore, the upper neck demonstrated relatively low geometrical uncertainty. However, deformation and position changes may be substantial in the middle and lower neck. In addition, the anterior part of the prophylactic CTV requires a smaller margin than the posterior part. The positional accuracy in the posterior neck region may result from large anatomical deformation, the fusion of the SCM with other nearby muscles that might mitigate accurate definition, and considerable shoulder position variation [32].

### Global registration vs local uncertainties

Advanced imaging guidance techniques allow for the correction of the target or OAR position before each IMRT session; thus, volume images, such as cone-beam or conventional CT, are widely used in routine practice [33]. Usually, the images acquired in peri-treatment are rigidly registered on the planning CT in the execution of image-guided IMRT. This rigid transformation does not account for all anatomical changes and most likely introduces more uncertainty during the alignment of the target volume or OARs. These inaccuracies cannot be neglected when the relative positions and shapes of the target volume and OARs change significantly [34]. Although deformable imaging registration enables local tracking of the target and organs and may harbor wide clinical application, its reliability warrants further evaluation [35].

Using weekly repeat CT images and rigid registration with bone matching to imitate radiation treatment in daily practice, we focused on the geometrical changes in the neck sublocations in this study. From the upper neck to the lower neck, the body contour decreased by 3.4–7.8% throughout the treatment course. Although the difference between the actual and registered planes was quite small (0.2–1.7%) in the middle and lower neck, the standard deviations were 3–5 times larger than their mean values, indicating considerable individual variability. The rigid registration and simple bone matching may have partially contributed to these inaccuracies. Even with online position correction and thermoplastic mask immobilization, the shoulder motion for patients with HNC was 2–5 mm in each direction on average [32] and the middle-lower neck displayed large body contour distortion. Finally, for some regions of the images, such as the hyoid bone and cricoid cartilage, matching is more difficult, resulting in more variable findings.

### Clinical applications

This is the first study, to the best of our knowledge, to demonstrate (Fig. 1C) the positional shift and quantified imaging registration-induced errors of the sublocations in the prophylactic neck CTV and estimated a required anisotropic CTV-PTV margin of 1–5 mm. Contouring each neck nodal level independently and defining an anisotropic margin in each direction would prove to be more challenging. However, we provided the framework to select more individualized margins. For example, our findings suggest that a more generous margin is needed when a positive lymph node appears in the middle-lower neck, especially at levels IV and V. Furthermore, image-guided IMRT could reduce setup uncertainties [36], however the geometrical changes of the target volumes and OARs could not be adapted only via the imaging matching. And an adaptive replanning strategy might be needed the re-delineation of the target volume and/or OARs and the resetting of the PTV margin [37].

Our study has several limitations. First, we estimated the geometrical margin but did not include dosimetric analysis, which will be investigated in future studies. Due to the interactions of the dose distribution near the PTV, the dosimetric margin may need to be smaller than the geometric margin [29]. However, the reference borders of the CTV have been shown to change during treatment, and the target volume may need to be updated in the mid-treatment for those who experienced considerable tumor shrinkage occurred [38]. Consequently, radiation planning based on a new target volume may have a different dose distribution. Second, the definition of sublocations might be affected by imaging quality, intra-observer variability, and contour subjectivity, among other factors. Third, these results should be interpreted with caution for other HNCs, as we focused specifically on NPC. The treatment response and radiation dose delivered to each neck nodal level may differ. Fourth, the delineation of CTVs in node-negative, node-positive, and postoperative necks is slightly different [17, 18]. For node-positive neck nodal levels, the probability of capsular rupture and extracapsular extension should be carefully considered, and it has been recommended that the CTV include the entire muscle [18], which might be different from the reference border, as with the anterior and posterior edge and deep surface of the SCM in this study. Finally, the geometrical changes of neck nodal levels VII–X [16] were not included in this study and require further investigation. Despite these disadvantages, our results are valuable for improving IMRT for NPC.

## CONCLUSION

During the course of IMRT for NPC, the lateral surface of the prophylactic neck CTV experience a global medial shift of < 1 mm. However, the geometrical changes demonstrated high sublocal variability, and the imaging registration-induced errors in the middle-lower neck were substantial. The middle-lower and posterior parts of the neck showed a larger positional shift than those in the upper and anterior parts of the neck. Even with weekly image-guided IMRT, the ideal geometrical PTV margin is anisotropic at 1–5 mm in each direction. The dosimetric margin and adaptive strategy warrant further investigation to improve IMRT for NPC.

## Data Availability

All data produced in the present study are available upon reasonable request to the authors

## Acknowledgement

We are grateful to Prof. Jan-Jakob Sonke from the Department of Radiation Oncology, The Netherlands Cancer Institute, Amsterdam, The Netherlands, who generously offered the research software (WRLDMATC) developed in-house for this research. We would like to thank Editage (www.editage.cn) for English language editing.

## Notes

**Conflicts of interest** All authors declare no conflicts of interest. This work was presented at the 60^th^ annual meeting of the American Society of Radiation Oncology, San Antonio, Texas, USA, October 21 - 24, 2018.

### Competing Interest Statement

The authors have declared no competing interest.

### Funding Statement

This study was funded by National Natural Science Funding of China (81974462).

### Author Declarations

This study was approved by the ethics committee of Hubei Cancer Hospital, Wuhan, China. And research data are stored in an institutional repository and will be shared upon request to the corresponding author.

